# Risks of Unregulated Home-Made Cannabis Products Used by Parents to Treat Refractory Epilepsy in Their Children

**DOI:** 10.1101/2025.04.25.25326163

**Authors:** Neubauer David, Žigon Dušan, Perković Benedik Mirjana, Bizjak Neli, Jasna Kovač, Matej Vehovc, Osredkar Damjan

## Abstract

**Introduction:** Some parents initiate treatment with non-prescription, home-made (unlabeled and unregulated) cannabis products for their children with refractory epilepsy without consulting their physicians. This study aimed to analyze the composition of these products in a pediatric cohort.

**Methods:** Home-made cannabis samples were analyzed for cannabinoid content, cannabidiol (CBD):Δ9-tetrahydrocannabinol (THC) ratio, and potential contaminants.

**Results:** Sixty home-made cannabis samples from 31 patients were analyzed. The most frequently detected cannabinoids were CBD, THC, and their acidic forms. The CBD:THC ratio varied significantly, with the most common ratio ranging from 1:1 to 10:1 (25.0%). Approximately 11.7% of the samples contained mainly CBD, 38.3% contained more THC than CBD, and 18.3% contained primarily THC with undetectable CBD levels.

**Conclusions:** Health professionals should strongly discourage the use of unregulated home-made cannabis products, as undisclosed and highly variable cannabinoid content with often elevated THC levels and potential contaminants present serious health risks for children with refractory epilepsy.

## Introduction

There is robust scientific evidence supporting the safe and effective use of cannabis-based products for treating epilepsy in select patient populations.(1) Specifically, adjunctive treatment with cannabidiol (CBD) has shown efficacy in reducing seizure frequency in children and adults with Dravet syndrome (DS) and Lennox-Gastaut syndrome (LGS) who experience seizures uncontrolled by standard anti-seizure medications (ASM).(2,3) Patients with other epilepsy syndromes, such as tuberous sclerosis complex, CDKL5 deficiency disorder, Aicardi, Dup15q, Doose syndromes, SYNGAP1 encephalopathy, and epilepsy with myoclonic absences, may also benefit from CBD as an adjunct therapy.(3) Our retrospective study indicated that CBD could significantly reduce the seizure burden in refractory childhood epilepsies of various causes, with 48.5% of patients experiencing more than a 50% reduction in seizures and 21.2% becoming seizure-free. Additionally, CBD treatment was associated with positive side effects, including improved behavior, better sleep, increased alertness, enhanced cognitive function, and improved speech.(4) CBD may also have a beneficial effect on quality of life (QOL) and mood independently of treatment response.(5)

Limited data are available on the safety of other cannabinoids and their impact on the seizure burden in patients with epilepsy. A report of a child with Dravet syndrome who had a significantly lower seizure frequency after treatment with full-spectrum cannabis extract has sparked much interest in the medical use of such products,(6) but their safety and efficacy have not been extensively studied in high-quality studies. The pharmacokinetics, efficacy in specific seizure/epilepsy types, dosing of the various cannabinoids, and adverse effects are still largely unknown.(7) Some studies suggest that when full-spectrum cannabis products are used, the average CBD dose may be lower than for products containing predominantly CBD, while achieving better reductions in seizure burden (71% vs 46%) and fewer adverse events.(8) A recent study on 5 pediatric drug-resistant epilepsy patients who failed management with an initial trial of high-dose CBD-focused therapy found there may be therapeutic benefit of adding THC to formulations.(9) Contrary to this, cannabinoids used for medical purposes in children and adolescents have also been linked to an increased risk of adverse effects in randomized controlled trials, such as diarrhea, elevated serum aspartate- and alanine aminotransferase levels, and somnolence.(10) Cannabis exposure is related to a higher seizure incidence than would be expected based on the prevalence of epilepsy in the general and pediatric populations; THC may be the causative xenobiotic for this phenomenon.(11) Furthermore, long-term cannabis usage is related to other medical problems such as cognitive deficits and smaller hippocampal volume in midlife.(12,13)

Despite increasing global access to legal pharmaceutical-grade cannabis formulations, many parents or caregivers of children with refractory epilepsy resort to non-prescription, unlabeled, and unregulated home-made cannabis products (sometimes also referred to as artisanal cannabis products) without seeking the advice of a physician. Limited data exists on the composition of nonprescription commercial cannabis products, but a study showed inconsistent labeling, deviations from their label claims should they exist, and lot-to-lot variability.(14) At the same time, cannabis-derived supplements can contain a complex phytochemical matrix.(15) The composition of home-made cannabis products is often undisclosed to consumers and remains poorly researched. An Australian study revealed significant variability in the cannabinoid content of cannabis extracts deemed “effective,” with no noticeable differences between those perceived as “effective” and “ineffective.” Contrary to families’ expectations, most products had low CBD levels, while THC was present in nearly every sample.(16) Further analysis of home-made cannabis samples from children with epilepsy showed that, although none exceeded heavy metal toxicity limits, 29% contained ethanol or isopropanol concentrations above generally accepted safety thresholds.(17) Other microbial contaminants such as fungi and bacteria can also be present.(18)

In Slovenia, physicians can prescribe the following medicines in the form of oral solutions which contain medical grade cannabis products to patients with pharmacoresistant epilepsy or cancer:

1. Pure medical grade CBD (THC-free) that contains 100 mg/ml of CBD.
2. Plant-based full-spectrum medical cannabis flower extract (17 mg/mL CBD/1.7 mg/mL THC), i.e. CBD:THC = 10:1.
3. Dronabinol (synthetic THC) that contains 10 mg/ml or 25 mg/ml of THC.
4. Medicines containing dronabinol and pure medical grade CBD in ratios 1:30, 1:20, 1:10, 1:2 and 1:1, where each ml contains 0.5 mg : 15 mg, 1 mg : 20 mg, 2 mg : 20 mg, 8.3 mg : 16.7 mg, and 8.3 mg : 8.3 mg, respectively.

However, the patients (their parents) in Slovenia have access also to food supplements and local/internet grey-market products containing cannabinoids. Labeling for content and concentrations of cannabinoids is often missing, inacurrate or misleading. For the unlabeled and uncontrolled products safety and efficacy data don’t exist. Products that were also often used by parents in Slovenia are produced by Charlotte’s Web.(6) For the latter, post-marketing surveillance reported low rates of adverse events (0.02%),(20) but their efficacy has not been evaluated in controlled clinical trials.

The objective of this observational study was to determine the composition of parent-administered home-made cannabis products, which the parents themselves obtained from unregulated sources without a physician’s consent, with a focus on the CBD:THC ratio of these products and the potential presence of contaminants.

## Methods

Parents obtained home-made cannabis products from unregulated sources and began administering treatment to their children before consulting their treating physician. Parents who disclosed the use of these products to physicians to treat refractory epilepsy or epileptic encephalopathy in their children between January and December 2022 were invited to participate in this observational study, conducted by the Department of Child, Adolescent, and Developmental Neurology at the University Children’s Hospital, University Medical Centre Ljubljana, Slovenia.

This observational questionnaire-based study was performed in accordance with the Declaration of Helsinki. This human study was approved by The National Medical Ethics Committee of the Republic of Slovenia - approval: 0120-131/2021/14. The results of the study can be found at https://repozitorij.uni-lj.si/IzpisGradiva.php?id=137168 (in Slovene). Parents were also invited to provide a sample of the substance they have used for the analysis and those who provided a verbal informed consent, participated in this analysis. Some provided more than one sample for the analysis during the study period; the products were from the same source, but not necessarily the same batch.

Investigated samples of cannabis products existed in a form of oils, resins, waxes or tinctures. Sample preparation involves weighing about 100 mg of sample on an analytical balance, extracting it in an ultrasonic bath in 1 ml of methanol, and diluting the extract tenfold, one hundredfold, and one thousandfold to 1 mL methanol solution.

Internal analytical method of liquid chromatograph coupled to mass spectrometer (LC-MS) were established for screening, identification and characterization of cannabinoids in samples. Mass spectrometer Q-Tof Premier (Micromass, Waters, Manchester, UK) coupled with Ultra performance liquid chromatograph (Acquity UPLC) system (Waters, USA) was used. The mobile phases consisted of 0.1% formic acid in LC-MS grade water (solution A) and organic solvent acetonitrile (solution B). All solvents and other chemicals were supplied by Sigma-Aldrich company. The elution gradient was linearly increased from 50% A to 95% B in seven minutes and go back to initial conditions at 8 min. The flow rate of mobile phase was 0.3 mL/min. An UPLC BEH-Phenil (1.7 µm, 15 cm x 2.1 mm internal diameter) chromatographic column was used. Autosampler and column oven temperature were set to 10 ^°^C and 40 ^°^C, respectively to prevent temperature degradation of acidic forms of cannabinoids. Injected volume of 1 to 5 µl samples were introduced through electrospray ionization source (ESI). A cone voltage of 20 V and a capillary voltage of 2.5 kV in negative ion mode (ESI-) and 3 kV in positive ion mode (ESI+), respectively were used. The desolvation gas of pure nitrogen was used at flow rate 600 L/h and desolvation temperature 350 ^°^C. ESI source temperature was set to 120 ^°^C. Mass spectra were acquired in centroid mode over an m/z range of 50-1000 in scan time 0.2 s and inter scan time 0.02 s. Data was obtained with MassLynx software version 4,1 (Waters, USA).

A mass resolution of approximately 9000 full widths at half maximum (FWHM) of the peak was applied for accurate high-resolution mass measurements (HRMS). The Leucin-Enkephalin solution in concentration 1 ng/ml was inserted parallel as lock mass to control mass accuracy of the Tof mass analyzer.

Accurate mass measurements define a unique elemental composition of protonated molecular ions (M+H)^+^ or deprotonated molecular ion (M-H)^-^ observed in mass spectra of LC-MS measurement of sample of products, such as, for example: m/z = 315.2329^+^ for C_21_H_31_O_2_ which corresponds to CBD or THC, or m/z = 357.2065^-^ for C_22_H_29_O_4_ which corresponds to CBDA or THCA within 3 ppm tolerance which is enough for confirmation of presence of cannabinoids in sample.

Quantitative determination of cannabinoids in samples based on calibration curves of standard methanol solutions of cannabinoids. Various analytical standards of cannabinoids were used: CBD standard 1 mg/mL methanol (Cerilliant Sigma-Aldrich company); Δ9THC standard solution 1 mg/mL methanol (Cerilliant Sigma-Aldrich company); Δ9THCA standard 1 mg/mL methanol (Cerilliant Sigma-Aldrich company); CBN standard 1 mg/mL methanol (Cerilliant Sigma-Aldrich company); standard mixture of three cannabinoids (CBD, Δ9THC and CBN) each at 1 mg/mL in methanol (Restek Accredited reference material producer).

Certified reference material: Cannabinoids Neutral 9 Standard (Restek Accredited reference material producer) in concentration 0.1% (1000 µg/mL) of each in P&T methanol which contain: Cannabicyclol (CBL), Cannabidiol (CBD), Cannabichromene (CBC), Delta-9-tetrahydrocannabinol (Δ9THC), Cannabigerol (CBG), Tetrahydrocannabivarin (THCV), Cannabinol (CBN), Delta-8-tetrahydrocannabinol (Δ 8THC), Cannabidivarin (CBDV).

The validation results indicated linearity of the LC-MS method in the concentration range of 0.01 to 1% (weight percentage) for all standards solutions, 0.003% limit of quantification (LOQ), 0.001% limit of detection (LOD) for most of detected cannabinoids, precision (inter/intra-day error less than 15% at fast chromatographic run time 8 min.

The concentration of cannabinoids in samples was calculated from peak area in LC-MS chromatograms. Identification and characterization of cannabinoids in samples were based on comparison of retention time of cannabinoids standards and compounds in samples, accurate mass measurements of protonated or deprotonated molecular ions in mass spectrum and elemental composition calculations of these ions measured with high resolution hybrid quadrupole orthogonal acceleration time-of-flight mass spectrometer (Q-Tof MS). With this analytical procedure some other organic compounds were also identified: terpenes, fatty acids, hormones etc.

In the samples of cannabis products were detected mostly cannabinoids: CBD and its acid variant (CBDA); 9-deltatetrahydrocannabinol (THC) and its acid variant (THCA); cannabinol (CBN); cannabichromene (CBC) and its acid variant (CBCA); cannabigerol (CBG) and its acid variant (CBGA); and cannabivarin (CBV) and its acid variant (CBVA). In the CBD:THC ratio calculations the total amounts of native and acidic forms of CBD or THC are summarized.

## Results

From the initial group of 51 patients whose parents disclosed the use of home-made cannabis products in their children, we excluded those with other diagnoses (such as autism spectrum disorders, Gilles de la Tourette syndrome, severe attention deficit disorder, and cerebral palsy). The final number of patients with a primary diagnosis of severe epilepsy/epileptic encephalopathy who received parent-administered home-made cannabis product was 31. Sixty samples of home-made cannabis products were obtained for chemical analysis.

The cannabis products were produced using different methods (the researchers had no information regarding the production process) and consisted of oils, waxes, resins, and tinctures. Among the 60 samples analyzed, CBD was detected in 49 (81.7%), CBDA in 45 (75.0%), THC in 54 (90.0%), THCA in 29 (48.3%), CBC in 38 (63.3%), CBCA in 4 (6.7%), CBG in 13 (21.7%), CBGA in 4 (6.7%), and CBN in 43 (71.7%) samples. All other cannabinoids were below the detection limit (weight percentage, < 0.01%).

The CBD:THC ratio varied significantly among the samples (Figure 1). The most frequent CBD:THC ratio in our samples was in the range of 1:1 to 10:1 (15/60 samples, 25.0%). Of all samples, 23/60 (38.3%) contained more THC than CBD, 2/60 (3.3%) contained CBD and THC at approximately equal concentrations, and 35/60 (58.3%) contained more CBD than THC (see Figure 1 for details). Interestingly, 7/60 (11.7%) samples contained only CBD, whereas THC was below the detection limit, and 11/60 (18.3%) samples contained only THC, whereas CBD was below the detection limit. In addition to CBD and THC, all samples contained lower concentrations of the other cannabinoids. In one sample, CBC and CNG were the predominant cannabinoids, whereas the concentrations of CBD and THC were below the detection limit.

**Figure 1.**
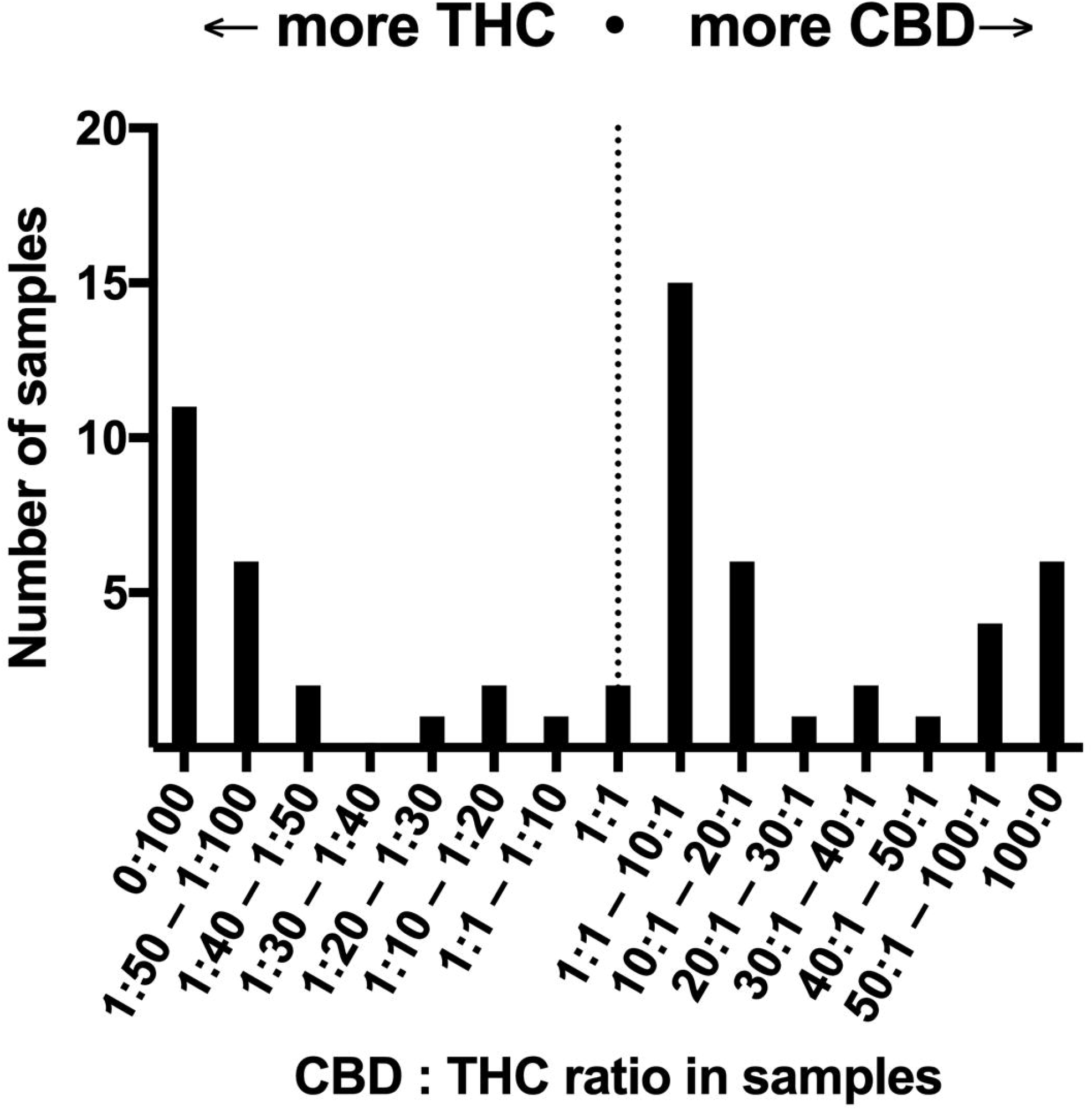
CBD:THC ratio of parent-initiated artisanal cannabis products for the treatment of refractory epilepsy in children

## Discussion

We anticipated some variability in the unlabeled home-made cannabis products caused by lack of quality control for these products, however, the study revealed unexpectedly high variability in composition profiles. While CBD is typically regarded as a beneficial component for treating refractory epilepsy, 38.3% of the samples contained more THC than CBD, and 18.3% contained primarily THC with undetectable CBD levels. These findings highlight the significant safety risks associated with home-made cannabis.

Refractory epilepsies present a substantial burden for both patients and their families.(21) When standard ASM and pure CBD preparations fail to sufficiently reduce seizures in refractory epilepsy, full-spectrum cannabis products may be considered a potential option by patients or parents, despite the lack of solid scientific evidence supporting their safety and efficacy. Some patients, including Slovene patients, turn to full-spectrum cannabis extracts available over local/internet gray markets. (6)(20)Uncontrolled quality and variability in the composition of home-made cannabis preparations pose significant risks to patients and previous studies have questioned their safety.(22) The LC-MS method has proven to be useful for testing the composition of these products. Our study found some home-made cannabis products with extremely high THC levels. Other studies have reported ethanol or isopropanol concentrations exceeding safety thresholds in these products.(17) Additionally, high THC levels in home-made cannabis products pose considerable risks to children, not only due to THC’s acute psychotropic effects but also its potential to exacerbate seizures or impair cognitive development.(11,13,23) For these reasons, the use of home-made cannabis products should be strongly discouraged. Physicians treating patients with refractory epilepsy should actively inquire about the use of these products as many patients may not disclose their use unless specifically asked.

One strategy to minimize or eliminate the use of home-made cannabis products by parents is to offer safe and regulated alternatives. While the regulatory frameworks for cannabinoids are still evolving, it is legally possible to prescribe and use such medicinal products in Europe.(24) In Slovenia, medical-grade cannabis products can be prescribed when certain conditions of the prescriber and the intended use are met: pediatric neurologists can prescribe various medical-grade magistral cannabis preparations to patients with refractory epilepsy as add-on therapy. These products are made by the Hospital pharmacy of the University Medical Centre Ljubljana and include pure CBD preparations, as well as preparations with controlled CBD:THC ratios of 10:1, 20:1, or 30:1 (with or without the presence of other cannabinoids at lower concentrations). All products were made from GMP-certified substances. Therefore, in Slovenia, whenever we encounter a pediatric patient whose parents have initiated treatment with home-made cannabis products, we strongly advise the parents to discontinue its use and potentially replace it with one of the medical-grade products available at our pharmacy.

## Conclusions

Parents of children with refractory epilepsy may turn to home-made cannabis products out of desperation or due to a belief in the benefits of “natural” treatments. However, our study revealed a high variability in the composition of these unregulated products. A significant number of samples contained very high THC levels. Healthcare professionals should strongly discourage the use of home-made cannabis products and recommend safer alternatives such as prescription medical-grade cannabis products. There is an unmet need for controlled, prospective studies to assess the safety and efficacy of products containing various cannabinoids, ideally comparing them to the more extensively studied CBD.

## Data Availability

All data produced in the present study are available upon reasonable request to the authors.

## List of abbreviations

CBC: cannabichromene
CBD: cannabidiol
CBG: cannabigerol
CBN: cannabinol
FDA US: Food and Drug Administration
THC: tetrahydrocannabinol
UPLC-MS: Ultra performance liquid chromatography-mass spectrometry

## Statements

### Ethical Approval and Consent to participate

This observational questionnaire-based study was performed in accordance with the Declaration of Helsinki. This human study was approved by The National Medical Ethics Committee of the Republic of Slovenia - approval: 0120-131/2021/14. The results of the study can be found at https://repozitorij.uni-lj.si/IzpisGradiva.php?id=137168 (in Slovene). Parents were also invited to provide a sample of the substance they have used for the analysis and those who provided a verbal informed consent, participated in this analysis.

### Consent for publication

All authors have consented to the publication of this article.

### Data Availability Statement

The data that support the findings of this study are not publicly available due to privacy reasons but are available from the corresponding author upon reasonable request.

### Authors’ contributions

ND: study design, evaluation of patients, analysis and interpretation of data, writing of the paper

ŽD: study design, analysis of blood samples

PBM: study design, evaluation of patients

BN: evaluation of patients, writing of the paper

JK: study design, evaluation of patients

MV: writing of the paper

OD: study design, analysis and interpretation of data, writing of the paper

## Acknowledgements

Not applicable

